# The real-world evidence to the effects of primary psychological healthcare system in diluting risks of suicide ideation in underrepresented children/adolescents: an observational, multi-center, population-based, and longitudinal study

**DOI:** 10.1101/2024.02.25.24303358

**Authors:** Wei Li, Xuerong Liu, Qianyu Zhang, Xiaobing Tian, Jidong Ren, Xiaodi Han, Chang Shen, Yanyan Li, Ji Chen, Lei Xia, Jingxuan Zhang, Yi Wu, Jie Gong, Hai Lan, Yan Wu, Psychological Health Guard for Children and Adolescents Project of China (CPHG) consortium, Chinese Open Science Network (COSN) consortium, Zhengzhi Feng, Zhiyi Chen

**Author notes:** Corresponding at: Zhiyi Chen or Zhengzhi Feng, Experimental Research Center for Medical and Psychological Science, School of Psychology, Third Military Medical University; Gao Tan-Yan Main Street, Shapingba, Chongqing, P.R. China. These authors contributed equally to this work (Wei Li, Xuerong Liu, Qianyu Zhang).

## Abstract

**Background:** Establishing primary psychological healthcare system to prevent suicide was eagerly advocated. However, it remains unclear whether such policy-driven and low-cost healthcare systems could be practical, especially with equal benefits for underrepresented children/adolescents. We aimed to examine the real-world practical effects of primary psychological healthcare system in preventing suicide ideation among children/adolescents, particularly underprivileged ones.

**Method and Findings:** The study employed an observational, multi-center, population-based and longitudinal design. 19,140 children/adolescents sampled from lower- and middle-income areas of western China (Nanchong) with 1-year followed-ups were included, of which majority to underrepresented underprivileged ones. The primary outcome was the incidence for reporting severe suicide ideation after practicing primary psychological healthcare system at 0.5-year and 1-year follow-ups, as contrasts to baseline. Subgroup analysis was conducted to examine the equal benefits of system for underrepresented children/adolescents. After instigating such system, the risks of reporting suicide ideation for included children/adolescents were found significantly lower compared with the control group at 0.5-year (adjust odds ratios [aOR] 0.28, 95%CI 0.23-0.33; p<0.001) and 1-year follow-ups (aOR 0.28, 95%CI 0.23-0.33; p<0.001). The effects were also observed among underrepresented children/adolescents including “left-behind” children/adolescents, “single-parent” children/adolescents and children/adolescents in especially difficult circumstance (CEDC, all pcorrected < 0.001). The effects in CEDC and “left-behind” children/adolescents were found non-inferior to typical developing cohort (all pcorrected < 0.01).

**Conclusions:** Primary psychological healthcare system was practically effective in reducing risks of suicide ideation in children/adolescents in 1-year, at least, but not yet with fully equal benefits for all the underprivileged ones.

## Introduction

Death by suicide is sharply becoming the one of the leading cause for increased mortality in the children/adolescents around the world.^1,2^ As official reports illustrated from the United Nations International Children’s Emergency Fund (UNICEF) in 2021, 4 per 100,000 children/adolescents aged at 10-19 died from suicide at home or even school per year.^3^ To make matters worse, given the prominently more risky factors derived from poor educational, healthcare or political resources, approximately 80% of suicide deaths occurred into low- and middle-income countries (LMICs) compared to advantaged ones.^4^ In China, almost 7 children/adolescents die from suicide per day, with 18-25% of this population having ever suicidal ideation. To control such suicide epidemics in children/adolescents, not only substantial financial budgets (e.g., Healthy China Initiative 2023, ¥1.1 billion) but also ambitious policy reform have been skewed into suicide prevention from establishing social psychological healthcare systems.^5-7^ Despite promising efforts and supports, the practices of these systems on preventing from suicide in children/adolescents still confronted challenging, as results of the fact that over 80% victims died from suicide even though they had ever reached out healthcare services.^8^ Thus, it is of imperative needs to explore practicable and effective psychological healthcare pattern for suicide prevention, especially for children/adolescents.

As one of the promising solutions for suicide prevention, there is a growing call from various sectors of society to integrate psychological services into the social primary healthcare system to ensure universally essential psychological health services with equitable accesses favoring to control suicide,^9-11^ but it still remains elusive for the practical roles of such changes. As official report drawn from the World Health Organization (WHO) in 2021, even though remarkable efforts have been made to establish and practice primary psychological healthcare system,^6^ one disappointing fact was that there still exits a worldwide failure to provide people in need with mental health services, due to the global shortfall in investment in mental health.^12^ Over 70% of total mental health expenditures in middle-income countries are allocated to for-profit hospitals/institutions, far exceeding investment in primary psychological healthcare system.^13^ Even more worryingly, the effectiveness of the current system is still questionable, especially in lower-middle-income western Pacific areas. Such non-profit healthcare system does not encourage high-quality services, leading to limited infrastructure, shortage of human resources and limited community awareness of psychological health.^14^ In addition of the “low-quality” concern brought by inadequate investment, primary psychological healthcare system still needs to address potential inaccessibility and unavailability for children/adolescents under underprivileged conditions^3^. Given that children/adolescents are under developmental for economic status and independent self-care ability, with thoughts and behaviors shaped and constrained by their parents/caregivers, their help-seeking intentions and actions usually arrested by their parents due to poor mental health literacy and suicide stigma especially in Chinese cultures.^15,16^ From what has been discussed for these existing challenges, it necessitates to provide real-world evidence clarifying the practical effects of such healthcare system as a policy for suicide prevention among the whole group of children/adolescents.

In contrast to primary physical healthcare system, the representation of children/adolescents, particularly those from underprivileged circumstances in LMICs, remains disproportionately negligible when practicing the primary psychological healthcare system, despite the principle of equitable distribution being the primary focus in the system. In LMICs, the number of psychiatrists specializing in treating children and adolescents was fewer than 0.1 per 100,000, significantly lower than 5.5 per 100,000 in high-income countries.^3^ Thus, the shortage of essential psychological services may handicap their children/adolescents in need from help-seeking. Unlike to the common sense for the needs of primary physical healthcare, stigma associated with mental health problems functions as a substantial societal and cultural barrier to the practices of primary psychological healthcare system among children/adolescents, especially in the lower-middle-income areas. As the results of less mental health knowledge and educations in such areas, the poor literacy leaves parents or even patients themselves to evaluate the mental problems as immoral and disgraceful “dirty disorder”.^17^ Several LMICs even legally criminate suicide behaviors, further increasing the their stigmatization for children/adolescents.^18^ On the other hand, children/adolescents living with underprivileged circumstances has long been underrepresented in the healthcare system. Compared with typical developing children/adolescents, underprivileged individuals living in disadvantaged socio-cultural environments (e.g., orphans, de facto unattended children/adolescents, “left-behind” children/adolescents, “single-parent” children/adolescents, and children/adolescents in especially difficult circumstances) experience greater psychiatric burdens and suicidal risks,^19^ but are more likely to “dropped out” from primary healthcare systems, as the fact that healthcare centers or service providers are geographically limited to economically developed urban areas, as well easily accessed for children/adolescents who are under well-parenting only.^20,21^ Therefore, we still need evidence to show whether underprivileged children/adolescents in LMICs equally benefit from psychological healthcare, examining whether such system provides equitable mental health rights int he real-world practices.

In June 2022, the Nanchong, one of biggest lower and middle-economic-status city including over 60% children/adolescents who are in underprivileged conditions in the western China, instigated a citywide primary psychological healthcare system (the Psychological Health Guard for Children and Adolescents Project of China, CPHG) for all eligible children/adolescents, especially in five underprivileged cohorts (i.e., children/Adolescents in especially difficult circumstance, CEDC, “Single-parenting” children/adolescents, “Left-behind” children/adolescents, de facto unattended children/adolescents and orphan). This system employed a “2+2” primary psychological healthcare pattern, including two-round psychological screening for risks of suicide and two-round psychological cares, The follow-ups for the incidences of suicide ideation were conducted at May. 21, 2023 and Oct. 29, 2023 for all the cohorts, respectively.

## Methods

### Study design and participants

A citywide, observational, multi-center, population-based, and longitudinal cohort study of children/adolescents (aged at 10-19) was conducted at Nanchong, Sichuan Province, China, to investigate the real-world effects of instigating primary psychological healthcare system on controlling the risks of suicidal ideation. The Nanchong was a representative lower-middle-economic-status city in the western China (National Bureau of Statistics of P.R.C., 2022). This study was implemented by the CPHG group.

We firstly established 385 healthcare centers in each middle and high school, as well as social welfare institutions in Nanchong. We assessed suicide ideation with a self-report signaling question at baseline, 0.5 year, and 1 year. All children/adolescents who reported suicide ideation in the past two weeks at baseline were not included in the final analysis, but they received immediately psychological services, clinical assessment, and medical treatment when necessarily. Thus, at the baseline, all the healthy children/adolescents were included in the current study.

Based on the locally legal definition to the underprivileged conditions (Ministry of Civil Affairs of the PRC, 2019), the enrolled children/adolescents in the present study were further categorized into five underprivileged cohorts, including de facto unattended children/adolescents, “left-behind” children/adolescents, “single-parent” children/adolescents, children/adolescents in especially difficult circumstance (CEDC), and orphan, as well as grouped for typically developing children/adolescents who were free from above conditions. The criterion of underprivileged adolescents can be found in supplementary materials (Appendix pp. 4).

This study is reported as per the Strengthening the Reporting of Observational Studies in Epidemiology (STROBE) guideline (Appendix pp. 14).

Informed consents were digitally signed by their parents or legal carers. This study has been formally approved by the Institutional Review Board (IRB) of The Sixth People’s Hospital of Nanchong (IRB-2022.002).

### Procedure

This system that the CPHG adopted was the “2+2” psychological healthcare pattern. The former “2” represented two-round psychological screenings for capturing the suicide ideation, In the first round, we used Center for Epidemiological Studies-Depression Scale (CES-D) to investigate whether children/adolescents reported depressive symptoms. The individuals with CES-D scores≥16 would be identified as “at-risk” children/adolescents for undergoing the second-round screening of directly measuring suicide ideation by a single signaling question (i.e., have you ever felt hopeless for the future, give rise to the idea of suicide?). Once identifying the suicide ideation, those ones would receive two-round specific psychological cares (interviews, the latter “2”). The first-round primary psychological care was carried out by trained and qualified staffs at healthcare centers following structural guidelines. A portion of them who were still identified “at-risk” for suicide ideation after primary psychological care would be transferred to the second-round preclinical (early) psychological interventions that conducted by clinicians at government-sponsored hospitals (Appendix pp. 7). Those children/adolescents would be transferred for inpatient medical treatments/interventions in case they were diagnosed for “high-risk” or clinically psychiatric disorders. Screening their depressive symptoms and suicide ideation has been implemented by the purpose-built online software in the cellphone. Meantime, healthcare centers lacking such infrastructures were supplemented by offline pencil-and-paper questionnaires. The practices of screenings were guided and supervised by CPHG staffs who trained and qualified ahead of this system implementation.

### Outcome

The primary outcomes were changes of incidence of reporting suicide ideation after instigating this primary psychological healthcare with “2+2” pattern at 0.5-year and 1-year follow-ups, compared to the ones at baseline.

### Statistical analysis

Descriptive statistical analyses were conducted using R (version 4.3.1), SPSS (IBM, Inc., version 29.0.1.0) and SPSSAU. The Mann-Whitney test and chi-square tests were carried out to examine the homogeneity between CPHG (child/adolescents enrolled in the “2+2” workflow) and control group. The frequency (rate) was calculated with 95% confidence interval (CI, estimated by Boostrapping method at n = 5, 000) to quantify the incidence of reporting suicide ideation.

As for the inferential statistics, the generalized linear mixed-effect models (GLMM) were built by *lmerTest* package in R, to test the effect size of instigating primary mental healthcare system on reducing risks of suicide ideation. A random effect for clustering of children/adolescents within regions was accounted to capture variability between groups. Missing data were dummy coded in Sociodemographic characteristics and removed in CES-D and suicide ideation records. For singular fit occurred in GLMM, we performed generalized linear model (GLM) to simply the analysis. The GLMM for outcome included fixed effects for CPHG group and control, including comparison of the whole group and comparison of the subgroups (e.g., CEDC, orphan, de facto unattended children/adolescents, “single-parent” children/adolescents, and “left-behind children/adolescents”), adjusted for centers, age, sex, offspring, family condition (excluded in the comparison of subgroups) and depressive symptoms. By building upon logit regression model, we calculated original and adjusted odds ratios (OR) and 95% CI for CPHG group compared with control group at 0.5-year and 1-year follow-ups, respectively. Also, the absolute risk difference, relative risk reduction for such changes were calculated.

To further examine the whether such benefits are equal effects in underprivileged children/adolescents, we used non-inferiority tests that implemented by SPSSAU. Given that no evidence-based non-inferiority boundary values that provided previously, we tentatively limited this liberal boundary value to 40% of incidence rate of typical developing individuals. We estimated the sample size by using normal approximation method, with a two-sided α of 0.05, a β of 0.10. The minimum sample size was found to be 2,034. Holm-Bonferroni correction was employed to adjust p values for the multiple comparisons.

For sensitivity analysis, we have redone main analyses mentioned above by re-enrolling from propensity score (PS)-matched samples that calculated by nearest neighbor matching algorithm. After estimations to PS, the 2,807 children/adolescents of CPHG group who individually matching to ones in the control group were selected out. Homogeneity of these population-based covariables (e.g., age, sex, offspring, and family condition) between CPHG and control group were verified in the post-hoc analyses. Sensitivity of the current study was evaluated for the replications of these findings between original sample and PS-match one.

### Role of the funding source

The study was funded by Civil-Affairs Fund of Sichuan. The funder of the study was not engaged in the study design, data collection and analysis, interpretation of result, report writing, and the decision of submitting the paper for publication.

## Results

A total of 180,006 children/adolescents geographically representing the entire Nanchong were enrolled in this primary psychological healthcare system, All the eligible children/adolescents (n = 19, 140) derived from 326 centers were included in the present study, with 13,527 children/adolescents for undergoing “2+2 pattern” primary psychological healthcare and 5,613 ones for receiving traditional social healthcare (Fig 1 for details). Demographic characteristics for these groups have been tabulated (Table 1). Details for the subgroups (i.e., underprivileged cohorts) can be found at the Table 2.

**Fig 1:**
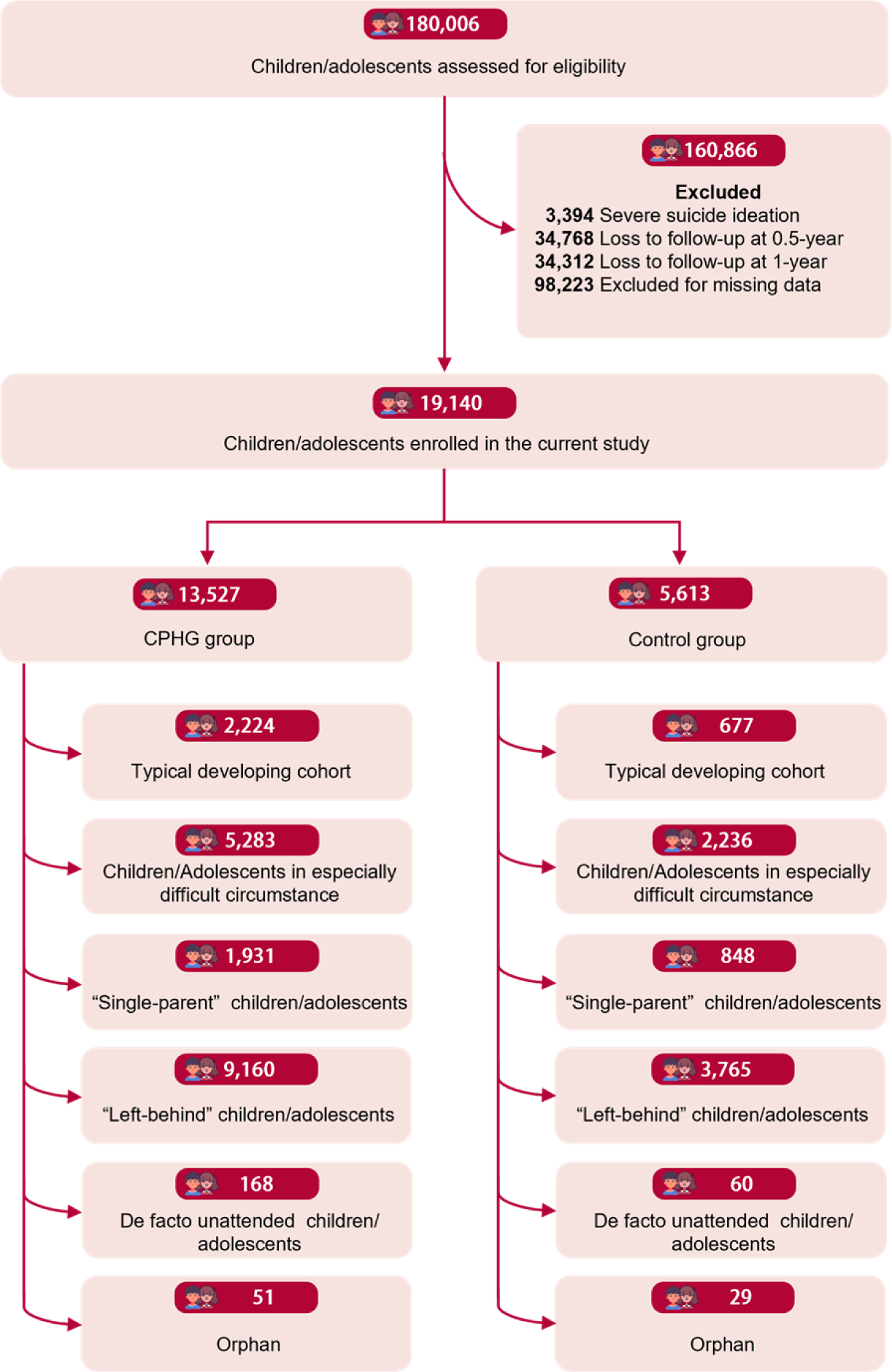
Screening and primary outcome population. The CPHG group refers to those who were enrolled in the Psychological Health Guard for Children and Adolescents Project of China (CPHG), whereas the control group was not enrolled in the CPHG.

**Table 1:** Sociodemographic characteristics of the population enrolled in this study. Data are mean (SD) or n (%). Missing=missing value. CES-D = Center for Epidemiological Survey, Depression scale. Data were extracted from CPHG group across 596 sites covering almost all the areas of Nanchong city (Sichuan, China). A total of 180,006 children/adolescents were included in CPHG group, whereas 19140 individuals were enrolled in the current study because of completely missing records, lack of informed consent, and loss to follow up. Underprivileged cohort consisted of orphan, de facto unattended children/adolescents, children/adolescents in especially difficult circumstance, “left-behind” children/adolescents, and “single-parent” children/adolescents. The criteria of these underprivileged conditions were detailed in supplementary materials (Appendix pp.4).

|  | Before PS matching |  |  |  | After PS matching |  |  |  |
| --- | --- | --- | --- | --- | --- | --- | --- | --- |
|  | CPHG | Control | Total | *p*-value | CPHG | Control | Total | *p*-value |
|  | (n=13527) | (n=5613) | (n=19140) |  | (n=2807) | (n=2807) | (n=5614) |  |
| **Age** | 14.22 (1.50) | 13.85 (1.48) | 14.11 (1.51) | <0.001 | 14.22 (1.49) | 14.22 (1.48) | 14.22 (1.48) | 0.998 |
| 10-11 | 75 (0.6%) | 62 (1.1%) | 137 (0.7%) |  | 11 (0.4%) | 10 (0.4%) | 21 (0.4%) |  |
| 12-14 | 7696 (56.9%) | 3673 (65.4%) | 11369 (59.4%) |  | 1616 (57.6%) | 1613 (57.5%) | 3229 (57.5%) |  |
| 15-17 | 5629 (41.6%) | 1854 (33.0%) | 7483 (39.1%) |  | 1159 (41.3%) | 1163 (41.4%) | 2322 (41.4%) |  |
| 18-19 | 127 (0.9%) | 24 (0.4%) | 151 (0.8%) |  | 21 (0.7%) | 21 (0.7%) | 42 (0.7%) |  |
| **Sex** |  |  |  |  |  |  |  |  |
| boy | 5146 (38.0%) | 2216 (39.5%) | 7362 (38.5%) | 0.77 | 1088 (38.8%) | 1081 (38.5%) | 2169 (38.6%) | 0.848 |
| girl | 8381 (62.0%) | 3397 (60.5%) | 11778 (61.5%) |  | 1719 (61.2%) | 1726 (61.5%) | 3445 (61.4%) |  |
| **Offspring** |  |  |  |  |  |  |  |  |
| Non-single child | 11259 (83.2%) | 4645 (82.8%) | 15904 (83.1%) | 0.42 | 2321 (82.7%) | 2330 (83.0%) | 4651 (82.9%) | 0.750 |
| Single child | 2268 (16.8%) | 968 (17.2%) | 3236 (16.9%) |  | 486 (17.3%) | 477 (17.0%) | 963 (17.1%) |  |
| **Family condition** |  |  |  |  |  |  |  |  |
| Typical developing cohort | 2224 (16.4%) | 677 (12.1%) | 2901 (15.2%) | <0.001 | 475 (16.9%) | 470 (16.7%) | 945 (16.8%) | 0.967 |
| Underprivileged cohort | 11210 (82.9%) | 4864 (86.7%) | 16074 (84.0%) |  | 2316 (82.5%) | 2322 (82.7%) | 4638 (82.6%) |  |
| Missing | 93 (0.8%) | 72 (1.3%) | 165 (0.9%) |  | 16 (0.6%) | 15 (0.5%) | 31 |  |
| **CES-D** | 24.40 (8.01) | 7.25 (5.03) | 19.37 (10.67) | <0.001 | 24.39 (8.08) | 7.54 (4.97) | 15.96 (10.77) | <0.001 |

**Table 2:**
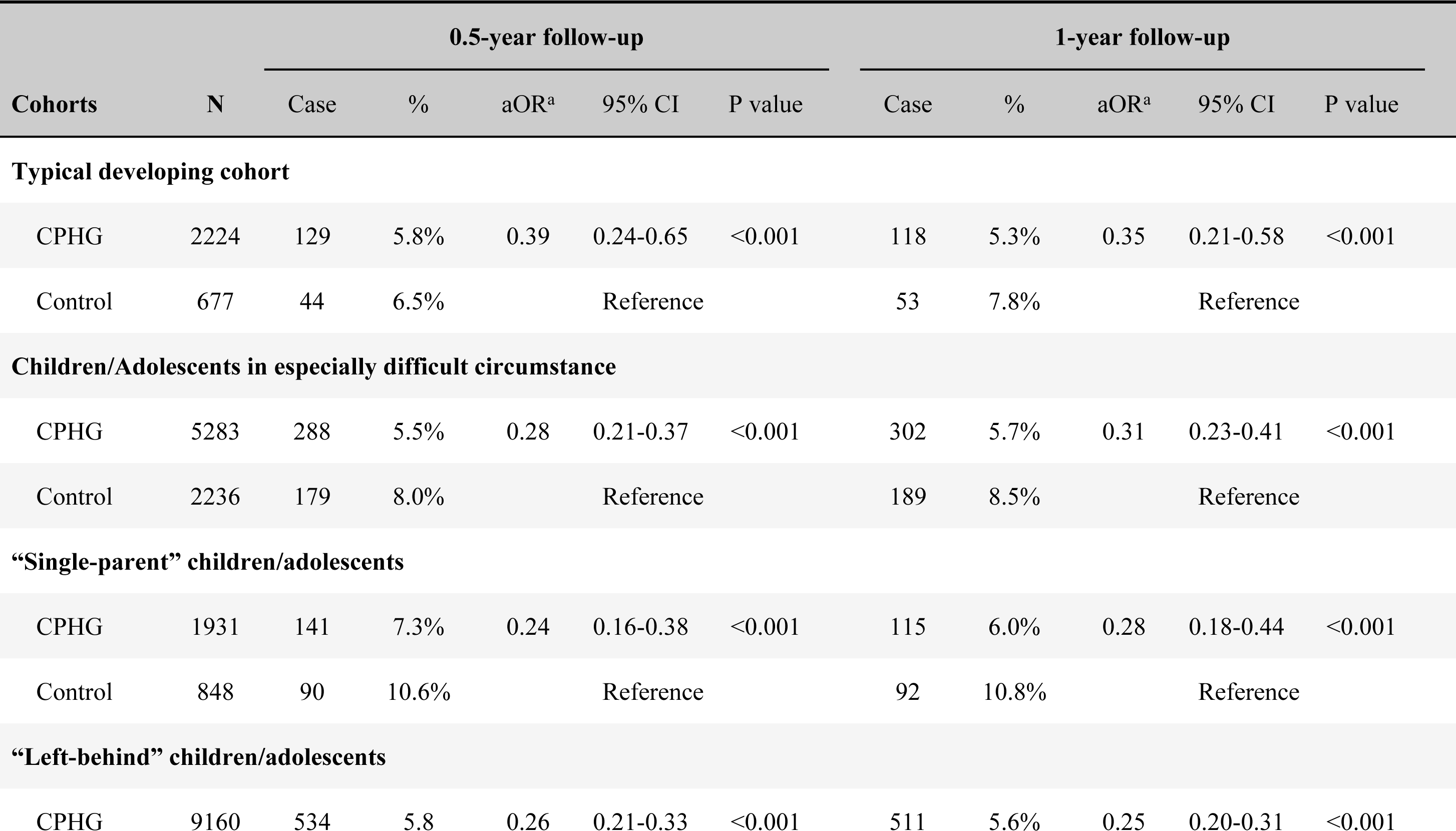
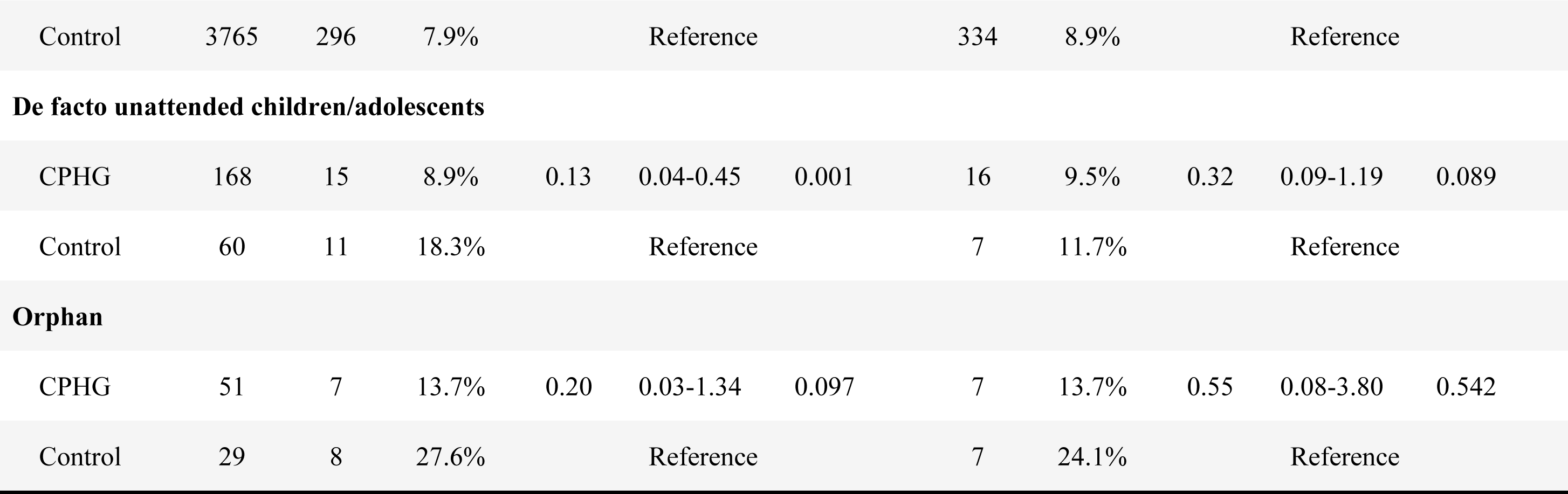
Subgroup analysis of the practical effects of implementing primary psychological healthcare system. Generalized linear mixed models were used for the analysis. a Adjusted Odd Ratios, adjusted for all Sociodemographic characteristics (age, sex, offspring) and depression estimated by Center for Epidemiological Survey, Depression scale (CES-D).

As shown in fig 2, at the 0.5-year follow-up, since instigating this “2+2 pattern” primary psychological healthcare system 5.7% (95% CI: 5.3 - 6.1, 772/13,527) children/adolescents in the CHPG group reported suicide ideation, whereas higher incidence reporting suicide ideation was found higher for children/adolescents in the control group (8.1%, 95% CI: 7.4 - 8.9, 475/5,613) Results of GLMM showed significant effects (adjusted OR = 0.28, 95% CI 0.23-0.33; p < 0.001) of practicing this system on reducing risks of reporting suicide ideation in the CPHG group, as contrast to ones in the control ones. Further, we observed the relative risk reduction (RRR) of 29.6% for the CPHG group compared to control ones (p < .01, Permutation test at n = 5,000; e.g., of 1000 children/adolescents, 57 ones reporting suicide ideation in the CPHG group *vs* 81 in the control group). At 1-year follow-up (fig 3), the incidence rate of reporting suicide ideation further decreased to 5.6% (95% CI: 5.3 - 6.0) in the CPHG group, but increasingly reached to 9.1% (95% CI 8.4 - 9.9) in the control group. The statistically significant 1-year-lasting effects (aOR = 0.28, 95% CI 0.23-0.33; p < 0.001, the same as 0.5-year follow-up) of implementing this system on reducing risks of reporting suicide ideation was still found in the CPHG group, as contrasts of control ones. The RRR was observed to be increasing to 38.5% (p < .01, Permutation test at n = 5,000).

**Fig 2:**
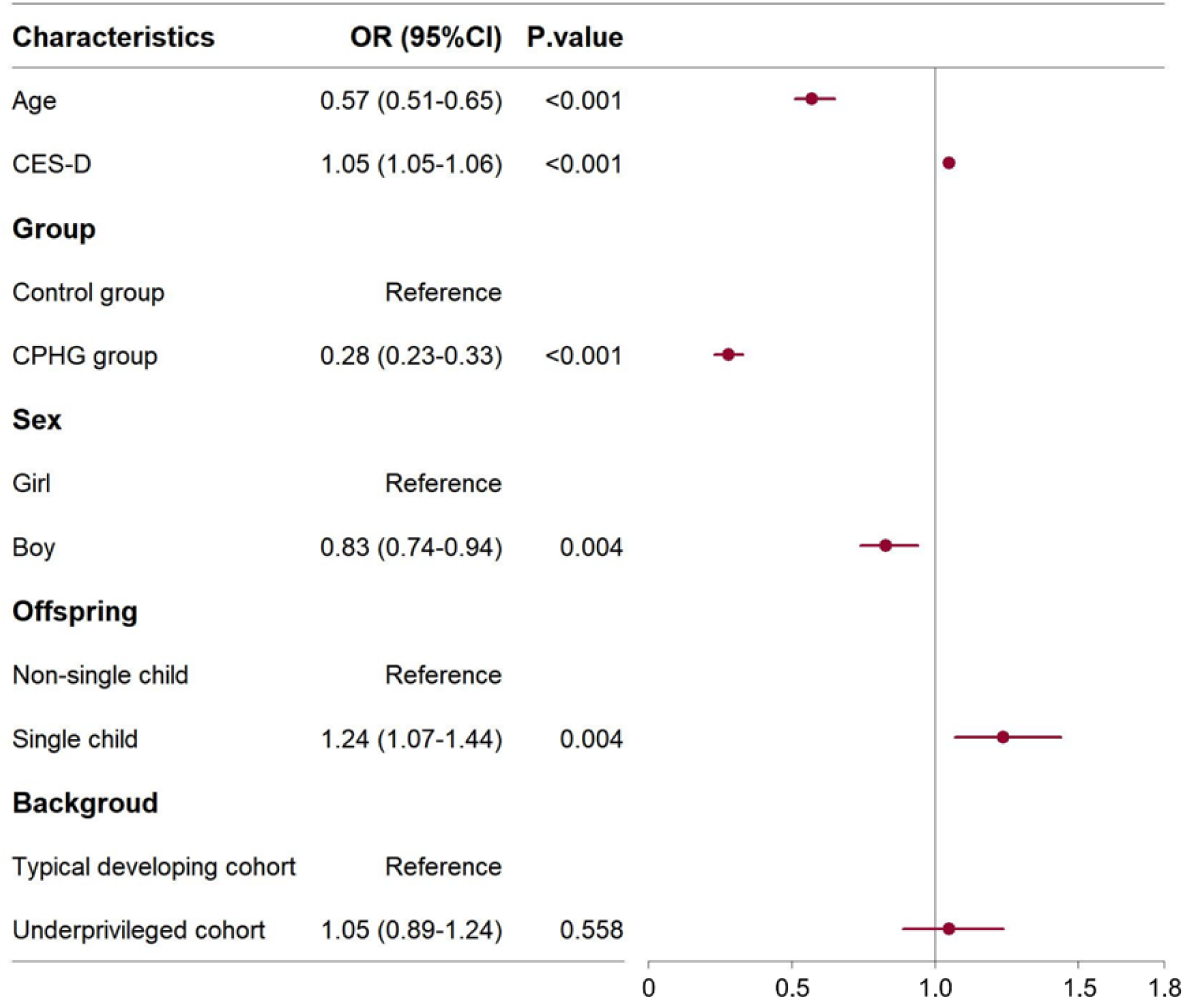
The Odd Ratio (OR) for the self-reporting suicide ideation at 0.5-year follow-up. The ORs were calculated by generalized linear mixed model. 95%CI=95% Confidence Interval. CES-D= Center for Epidemiological Survey, Depression scale. The CPHG group refers to those who were enrolled in the Psychological Health Guard for Children and Adolescents Project of China (CPHG), whereas the control group was not enrolled in the CPHG. When OR<1, the variable is considered as protective factor for preventing suicide ideation. When OR>1, the variable is considered as risk factor.

**Fig 3:**
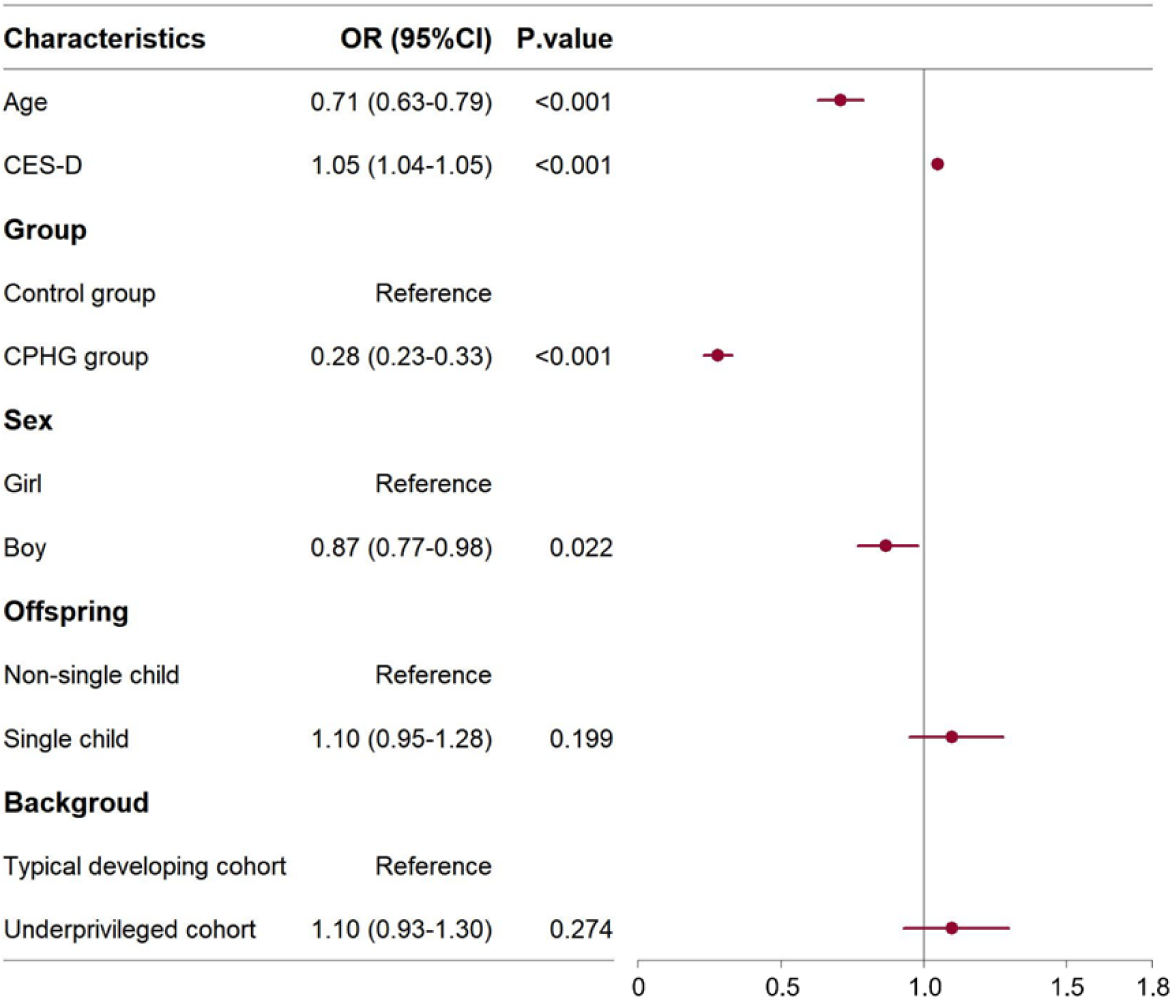
The Odd Ratio (OR) for the self-reporting suicide ideation at 1-year follow-up. The ORs were calculated by generalized linear mixed model. 95%CI = 95% Confidence Interval. CES-D = Center for Epidemiological Survey, Depression scale. The CPHG group refers to those who were enrolled in the Psychological Health Guard for Children and Adolescents Project of China (CPHG), whereas the control group was not enrolled in the CPHG. When OR<1, the variable is considered as protective factor for preventing suicide ideation. When OR>1, the variable is considered as risk factor.

In the subgroup analysis, at 0.5-year follow-up (May. 21, 2023), we found the statistically lower risks of reporting suicide ideation after practicing this “2+2 pattern” primary psychological healthcare, in not only typical developing children/adolescents (adjusted OR 0.39, 95%CI 0.24-0.65), but also these underprivileged cohorts including CEDC (adjusted OR 0.28, 95%CI 0.21-0.37), “single-parent” children/adolescents (adjusted OR 0.24, 95%CI 0.16-0.38), “left-behind” children/adolescents (adjusted OR 0.26, 95%CI 0.21-0.33) and de facto unattended children/adolescents (adjusted OR 0.13, 95%CI 0.04-0.45), as contrasts of control group (Table 2; all p ≤ 0.001, Benjamini-Hochberg corrections). At 1-year follow-up (Oct. 29, 2023), such benefits were observed in typical developing cohort (adjusted OR 0.35, 95%CI 0.21-0.58), CEDC (adjusted OR 0.31, 95%CI 0.23-0.41), “single-parent” children/adolescents (adjusted OR 0.28, 95%CI 0.18-0.44) and “left-behind” children/adolescents (adjusted OR 0.25, 95%CI 0.20-0.31) only (all p < .001, Benjamini-Hochberg corrections; Table 2).

Despite such prominent and (1-year) long-term benefits, by using non-inferiority tests, we further examined whether this system unexpectedly incurs inequality of such primary health benefits in underprivileged children/adolescents, including the CECD, “left-behind” and “single-parent” children/adolescents’ group, respectively, given their sample sizes reached statistical prerequisite. Results demonstrated the statistically non-inferior effects of practicing this system to CEDC and “left-behind” children/adolescents, compared to typical developing ones, irrespective of in 0.5-year and 1-year follow-ups (all p_corrected_ ≤ 0.01, Table S4). Exploratory analysis for the non-inferiority to other underprivileged cohorts can be found at Table S4.

As for the sensitivity analysis, we replicated main analyses by re-selecting PS-matched control group. Results were found to be parallel with main findings by showing the decreased incidence and risks of reporting suicide ideation after instigating primary psychological healthcare system in CPHG groups, compared to control group at both 0.5-year and 1-year follow-ups (Fig 4-5).

**Fig 4:**
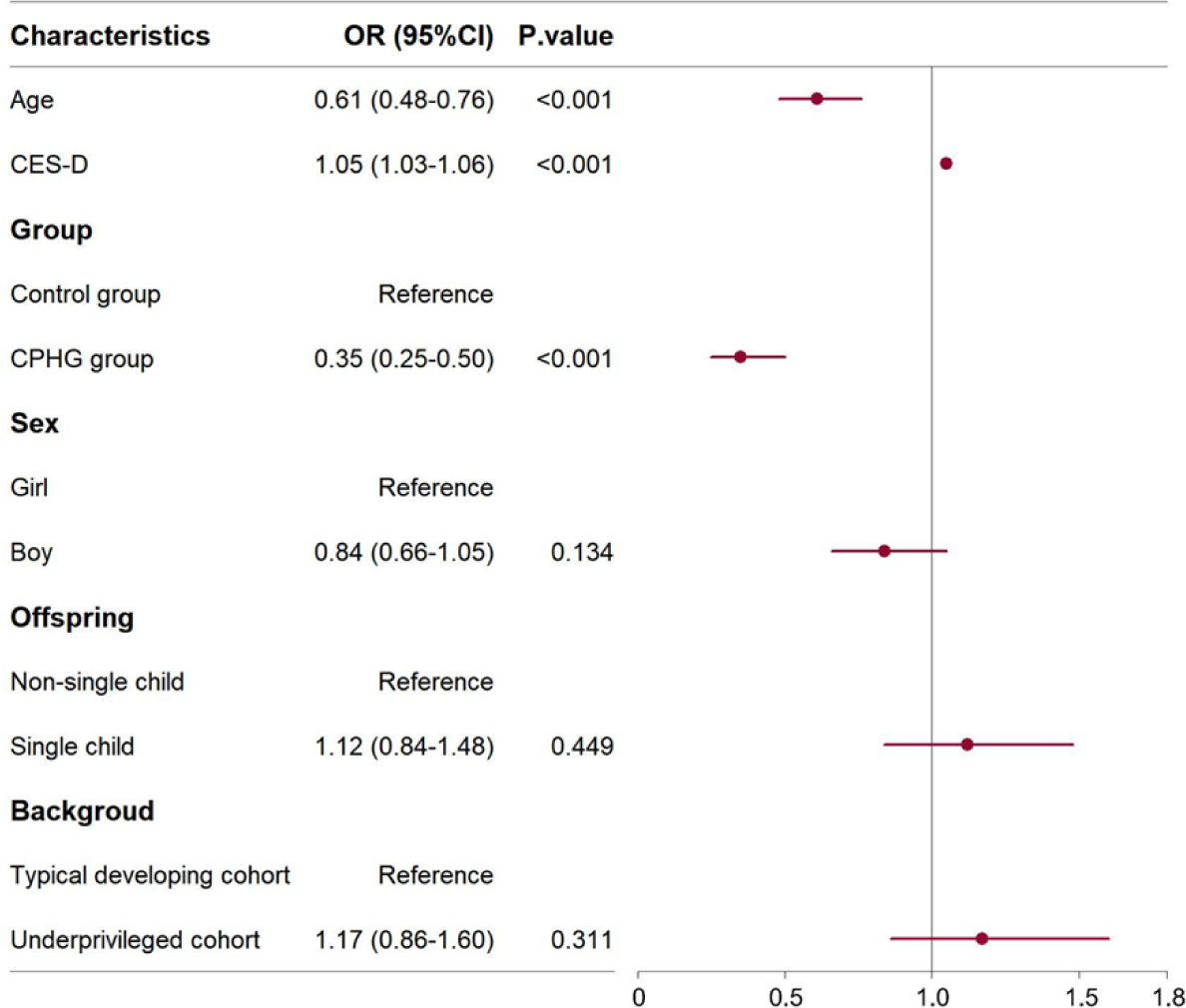
The Odd Ratio (OR) for the self-reporting suicide ideation after PS matching at 0.5-year follow-up. The ORs were calculated by generalized linear mixed model. 95%CI=95% Confidence Interval. PS matching = propensity score matching. The matching characteristics included age, sex, family condition, and offspring. CES-D = Center for Epidemiological Survey, Depression scale. The CPHG group refers to those who were enrolled in the Psychological Health Guard for Children and Adolescents Project of China (CPHG), whereas the control group was not enrolled in the CPHG. When OR<1, the variable is considered as protective factor for preventing suicide ideation, whereas the variable is considered as risk factor when OR>1.

**Fig 5:**
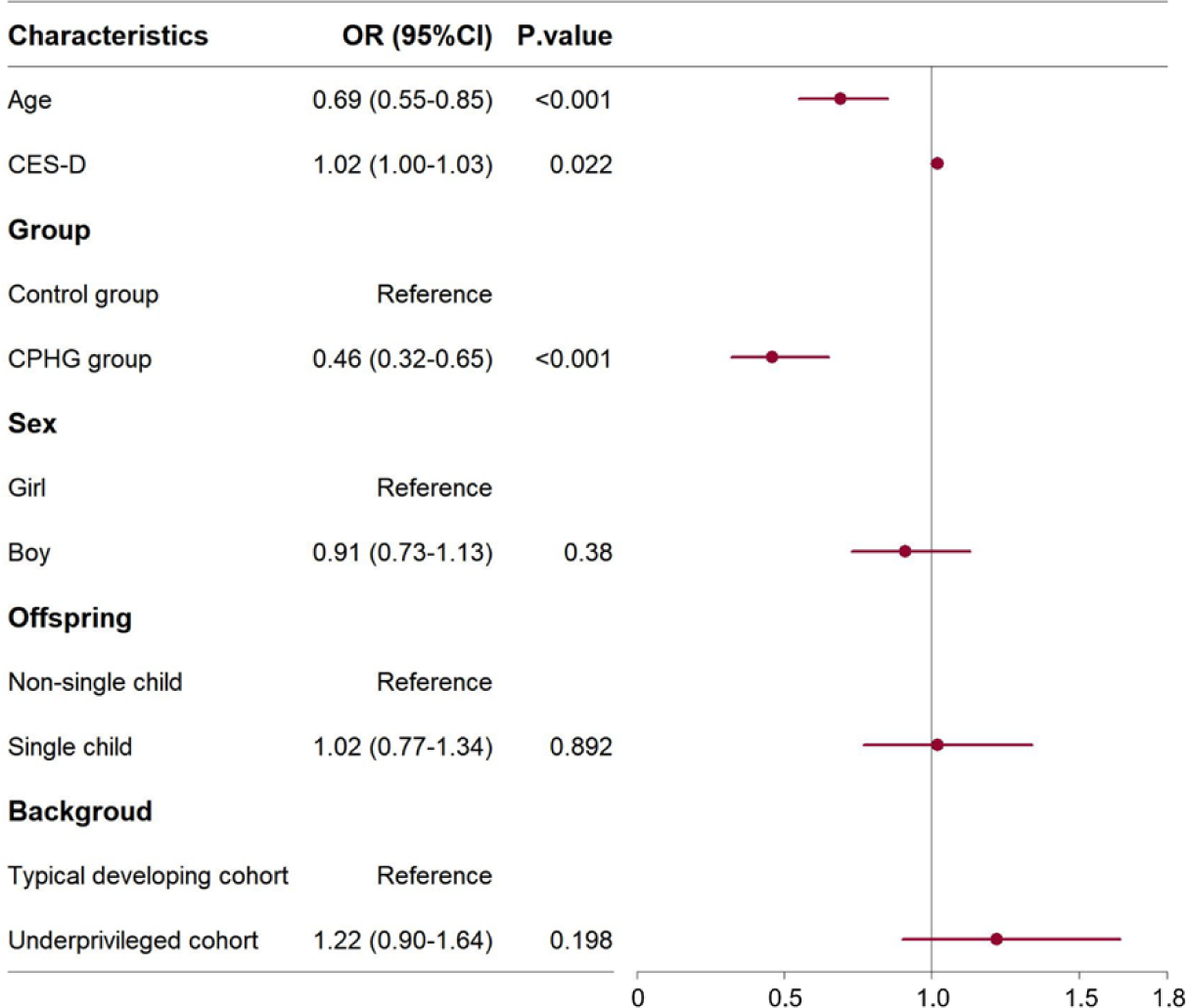
The Odd Ratio (OR) for the self-reporting suicide ideation after PS matching at 1-year follow-up. The ORs were calculated by generalized linear mixed model. 95%CI=95% Confidence Interval. PS matching = propensity score matching. The matching characteristics included age, sex, family condition, and offspring. CES-D = Center for Epidemiological Survey, Depression scale. The CPHG group refers to those who were enrolled in the Psychological Health Guard for Children and Adolescents Project of China (CPHG), whereas the control group was not enrolled in the CPHG. When OR<1, the variable is considered as protective factor for preventing suicide ideation, whereas the variable is considered as risk factor when OR>1.

## Discussion

This study provides the real-world evidence to illustrate the practical effects of establishing primary psychological healthcare system on reducing the risks of children and adolescents’ suicide ideation in a lower- and middle-income city. By implementing such “2+2 pattern” of primary psychological healthcare system, the reported reduction of relative risk was nearly 30% than for the control group at 0.5-year follow-up and nearly 40% at 1-year follow-up. In terms of the risks of psychological health inequality from this system, we found the absolute and relative risk reductions in not only the typical developing cohort but also all the underprivileged subgroups, including CEDC, “left-behind” children/adolescents, and “single-parent” children/adolescents, whereas failed to equally benefit to orphan and unattended children/adolescents in our exploratory analysis. Together, these results substantiated the real-world and (1-year) long-term benefits of establishing this citywide, low-cost, and population-based “2+2 pattern” primary psychological healthcare system to reduce risks of suicide ideation in children/adolescents living in lower-middle-income areas, even if under several specific underprivileged conditions.

The current study firstly examined the real-world effects of establishing a primary psychological healthcare system to control the risks of children and adolescents’ suicide ideation in a lower-middle-income city, and found nearly 40% relative risk reduction after implementing this system at even 1-year follow-up with a consistent adjusted OR. Although integrating primary healthcare and psychological services is considered as one of the most viable options for preventing and solving children and adolescents’ suicide issues,^21^ the role of such healthcare system in preventing suicide ideation among children/adolescent is still unclear so far. The supporting evidence drawn from a global meta-analysis reported the significantly decreasing in suicide ideation after integrating psychological healthcare into the primary system which employed a post-primary school-based suicide prevention (PSSP) pattern including awareness programs, screening, and interventions for high-risk children/adolescents.^22^ However, the contradictory results also illustrated null effects of integrative psychological services neither on controlling the incidence of suicide attempts or severe suicide ideation at follow-ups (e.g., The Screening by Professionals program),^23^ leading to the concern that such integrative primary psychological services may be not well enough to tackle “suicide epidemics”, especially for global children/adolescents. Therefore, the overarching and major contribution of the current study was to provide real-world evidence supporting the practical effects of establishing pattern-specific primary psychological healthcare on reducing the risks of children and adolescents’ suicide ideation.

The practical effects of implementing primary psychological healthcare system in the current study could be attributed to its rectification of the following deficiencies. On the one hand, because of limited mental health resources (licensed psychiatrists, psychiatric registrars, and psychiatric nurses) and millions of community-dwelling individuals in-need, the accessibility of psychological healthcare and medical services is still insufficient,^24^ despite the excessive mental health expenditures in the mental health, especially in LMICs.^13^ Therefore, the “2+2” psychological pattern combined with large-scale, multi-center, and population-based design in the primary psychological healthcare system could directly ameliorate the universally accessibility of early identifying of suicide ideation and early psychological services, covering all children/adolescents in need.^25^ On the other hand, stigma associated with suicide related issues among children and adolescents who are suffering from suicidal ideation handicap them from seeking psychological support, leading to more serious outcomes (suicide attempt and suicide).^26^ These children/adolescents who arrested by stigma are usually lack of mental health education and literacy as well as social support.^27^ Hence, despite that primary psychological healthcare system is categorized as low-cost project with “low-quality” concern, such brief psychological healthcare in this system could give them social support, enhance the mental health literacy about suicide related issues of children/adolescents, and help them to overcome the suicide crisis.^28^

Another finding worthy to be discussed is to specifically evaluate the potential risks of psychological health inequality from instigating this primary psychological healthcare system for underprivileged children/adolescents who are generally underrepresented in the family, school, and social cares. However, we observed a potentially moderate “inequality” for condition-specific underprivileged cohorts from this system, especially in unattended and orphan children/adolescents. Unlike to children/adolescents under parental or family-based cares, on the one hand, these unattended and orphan children/adolescents are confront with more mental health challenges from prominently lower socioeconomic positions, peer bullying and relevant disadvantageous conditions (e.g., poverty, less caregiving and lacking access to education), leading to higher risks for mental health problems than do of their peers.^29^ On the other hand, previous practices linking to primary healthcare services for children/adolescents, are highly dependent from full-fledged social systems (e.g., family, school, community), but are hard to individually reach unattended ones outside these systems.^30^ Given the worse benefits of this system for these unattended ones, we may prompt a urgent calls for specific psychological healthcare towards these underprivileged children/adolescents without well-established parenting cares. More importantly, generalizing this system elsewhere may need to design specific solicitude for these underrepresented cohorts.^31^

Despite merits, the several limitations in the current study should be considered These results were derived from observational cohorts in the policy changes to establish primary psychological healthcare (i.e., CPHG), but not yet from standard Randomized Controlled Trial (RCT) providing “casual and conclusive” evidences. In addition, given the cost-saving design for this universally primary healthcare system, we indeed lacked clear evidences to validate the healthcare quality (e.g., psychological cares, psychological screenings, individual evaluations to this “2+2 pattern” practices). Lastly, these data in the current study were drawn from a citywide real-world population, leading to unpredictable losses in the follow-ups, such as lacking of parental consents and cooperation.

In conclusion, the current study provided this empirical real-world evidence of the benefits of the population-based and low-cost primary psychological healthcare system to reducing citywide risks of suicide ideation for children/adolescents, especially in such lower-middle-income areas. What’s more, we found nearly 30% relative risk reduction after establishing this system at 0.5-year follow-up, and even observed 1-year long-term benefits by showing nearly 40% at 1-year follow-up. On the other hand, the current study highlighted the risks of health inequality for the benefits of this system on underrepresented children/adolescents, especially for ones in underprivileged parenting cares. Taken together, by conducting this observational, population-based and 1-year longitudinal study, we provided real-world evidences to substantiate the benefits of instigating the primary psychological healthcare system with “2+2 pattern” to reducing risks of children and adolescents’ suicide ideation, particularly in such lower-middle-income areas encompassing underrepresented ones who were in underprivileged conditions.

## Data Availability

All the follow-up data had been deposited into the Science Data Bank (ScienceDB, https://doi.org/10.57760/sciencedb.12150) for full accesses once being approved by the Data Regulation Office (CPHG-DRO). Form for applying the case-by-case approval could be found at the FigShare (https://doi.org/10.6084/m9.figshare.24297532.v1). All the analyses were implemented by the commercial software (i.e., SPSS, IBM.Inc.) and open-source R packages (e.g., MICE). All code used in R software had been deposited into the Open Science Framework (OSF, https://osf.io/e5h3a/) for full accesses.

## Consortia authorship

**For the Psychological Health Guard for Children and Adolescents Project of China (CPHG) Group**

Yue-Guang Liu,^4^ Xian-Yong An,^4^ Ji-Dong Ren,^4^ Xiang Yuan,^4^ Yi Zhang,^4^ Jian Yang,^4^ Wan-Xia Li,^4^ Yao Peng,^4^ Yao Peng,^4^ Yu-Jia Liao,^4^ Si-Yu Cheng,^4^ Lu Liu,^4^ Hong-Jiang Yu,^4^ Tian-Tian Liu,^4^ Li-Jun Liang,^4^ Meng-Zhu Cheng,^4^ Xi Zhao,^4^ Xiang-Yu Deng,^9^ Hui-Lei Xu,^9^ Jidong Ren,^4^ Xiaobing Tian,^3,4^ Xue-Hua Li,^4^ Yi-Han Wen,^4^ Jun Lei,^4^ Xiao He,^4^ Hong-Ying Liu,^4^ Lei Zhang,^4^ Zhen-Mi Liu,^9^ Xian-Dong Meng,^10^ Xia Jiang,^10^ Yuan-Yuan Li,^10^ Jiajun Xu,^10^ Zhengzhi Feng,^1^ Zhiyi Chen^1^

^9^ Department of Children and Maternal Health, Western China Hospital, Sichuan University, Chengdu, Sichuan 610065

^10^ Mental Health Center, Western China Hospital, Sichuan University, Chengdu, Sichuan 610065

## Contributors

Conceptualization, C.Z.Y., F.Z.Z., and L.W; Data curation, L.Y.Y., S.C., G.J., H.X.D., W.Y., and X.L; Formal analysis, L.W., L.X.R., and Z.Q.Y.; Visualization, L.W., L.X.R., and Z.Q.Y.; Funding acquisition, C.Z.Y. and F.Z.Z.; Project administration, C.Z.Y. and F.Z.Z; Resources, L.Y.G., A.X.Y., R.J.D., Y.X., Z.Y., Y.J., P.Y., L.Y.J., C.S.Y., L.L., Y.H.J., L.T.T., L.L.J., C.M.Z., Z.X., D.X.Y., X.H.L., L.X.H., W.Y.H., L.J., H.X., L.H.Y., Z.L., L.Z.M., M.X.D., J.X., L.Y.Y. and X.J.J.; Writing-original draft, L.W., L.X.R., and Z.Q.Y.; Writing-review and editing, C.Z.Y., F.Z.Z., C.J., T.X.B., Z.J.X., L.H., W.Y., and R.J.D. All authors have read and approved the publication of this manuscript.

## Declaration of interests

T.X.B., L.Z.M., M.X.D., J.X., L.Y.Y., X.J.J and G.J. disclosed to receive financial supports from this project. All other authors disclosed no competing interests.

## Acknowledgement

This study was conducted with the support of funding from the Civil Affairs Department of the Sichuan Government (China) and the Minors Protection Care Foundation of the Sichuan Charity Federation (China). We do appreciate all the efforts and contributions of the government and other sectors for children and adolescents’ mental health.

## Notes

### Competing Interest Statement

The authors have declared no competing interest.

### Funding Statement

Yes

### Author Declarations

This study has been formally approved by the Institutional Review Board (IRB) of The Sixth People’s Hospital of Nanchong (IRB-2022.002).

